# E-cigarette use and Respiratory Symptoms in Residents of the United States: A BRFSS Report

**DOI:** 10.1101/2022.05.30.22275770

**Authors:** Marcia H. Varella, Olyn A. Andrade, Sydney M. Shaffer, Grettel Castro, Pura Rodriguez, Noël C. Barengo, Juan M. Acuna

**Author notes:** These authors contributed equally to this work (Marcia H. Varella, Noël C. Barengo, Juan M. Acuna). These authors also contributed equally to this work (Olyn A. Andrade, Sydney M. Shaffer, Grettel Castro, Pura Rodriguez).

## Abstract

**Purpose:** E-cigarettes are the most common type of electronic nicotine delivery system in the United States, rapidly gaining presence worldwide. E-cigarettes contain numerous toxic compounds that have been shown to induce mild to severe structural damage to the airways. The objective of this study is to assess if there is an association between e-cigarette use and respiratory symptoms in adults in the US as reported in the BRFSS in an attempt to drive potential early detection and prevention of severe or permanent respiratory disease.

**Methods:** We analyzed data from 18,079 adults, 18-44 years, who participated at the Behavioral Risk Factor Surveillance System (BRFSS) in the year 2017. E-cigarette smoking status was categorized as current everyday user, current some days user, former smoker, and never smoker. The frequency of any respiratory symptoms (cough, phlegm, or shortness of breath) was compared. We calculated unadjusted and adjusted odds ratios (OR) and 95% confidence intervals (CI).

**Results:** The BRFSS reported prevalence of smoking e-cigarettes was 6%. About 28% of the participants reported any of the respiratory symptoms assessed. The frequency of reported respiratory symptoms was highest among current some days e-cigarette users (45%). After adjusting for selected participant’s demographic, socio-economic, and behavioral characteristics, and asthma and COPD status, the odds of reporting respiratory symptoms increased by 49% among those who use e-cigarettes some days (OR 1.49; 95% CI: 1.06-2.11), and by 29% among those who were former users (OR 1.29; 95% CI: 1.07-1.55) compared with those who never used e-cigarettes. No statistically significant association was found for those who used e-cigarettes every day (OR 1.41; 95% CI 0.96-2.08)

**Conclusion:** E-cigarettes cannot be considered safe alternatives to combustible traditional cigarettes. Cohort studies may shed more evidence on the association between e-cigarette use and specific respiratory diseases.

## Introduction

E-cigarettes use, or vaping was considered a safer alternative to conventional cigarettes and a potential alternative in tobacco smoking cessation efforts (1-3). It was introduced in the US marketplace in 2007 and since then its use has grown exponentially. Estimates based on the 2016-2017 Behavioral risk Factor Surveillance System (BRFSS) indicate a 4.4% prevalence of e-cigarette use for adults (18 years or older) (4,5) According to the National Youth Tobacco Survey (NYTS), e-cigarette use in 2020 for middle and high school students was estimated to be about 19.6 % and 4.7%, respectively (6).

The increase in the use of e-cigarettes is of concern. While e-cigarettes may reduce regular cigarette use, e-cigarette aerosol contains several toxic chemical constituents, including diacetyl or benzaldehyde, that negatively affect the respiratory system. (7,8). E-cigarette is also associated with wheezing, chronic cough, phlegm, or bronchitis in children and adults. (9-11). The results of a recently published systematic review estimated that the pooled OR associated with e-cigarette use for asthma was 1.39 (95% confidence interval (CI) 1.28-1.51) and for COPD was 1.49 (95% CI 1.36-1.65) (12).

While growing evidence points to detrimental respiratory effects of e-cigarettes use, just a few comparative studies assessed the effects of e-cigarettes in population-based, community-dwelling adults in the US(13-15). Additionally, the impact of e-cigarettes on the respiratory system has been assessed mainly by the report of asthma and or COPD, which could have underestimated the magnitude of the respiratory effects caused by e-cigarettes.

In this study, we assessed the association between e-cigarette use and the occurrence of respiratory symptoms (namely cough, phlegm production, or shortness of breath) as reported in the BRFSS, to obtain a more comprehensive understanding of the previous potentially underestimated impact of e-cigarette use in adults in the United States.

## Methods

### Design and setting

We used data from the 2017 BRFSS. Briefly, the BRFSS is an ongoing national yearly cross-sectional survey, originally aimed to identify emerging health problems to modify public health programs and policies. (16) It consists of phone-based interviews regarding participants’ health-related risk behaviors, chronic health conditions, and use of preventive services. The inclusion criteria consisted of individuals between the ages of 18-44 and those living within Arizona, the District of Columbia, Florida, Georgia, Minnesota, Nevada, Tennessee, and West Virginia. Only these states in the US collected information on the outcome of interest.

### Study variables

E-cigarette usage (or vaping) was categorized as current everyday e-cigarette smokers, current someday e-cigarette smokers, former smokers, and those that never smoked, according to self-report. The outcome of interest was the presence of respiratory symptoms, considered present if the participant reported any of the following: cough, phlegm production occurring daily in the past three months, or shortness of breath hurrying on level ground or when walking up a slight hill or stairs. No report of these three symptoms was considered as an absence of the outcome. Covariates assessed included participant’s age (assessed as a continuous variable and also categorized as either 18-34 or 35-44 years old), sex, race (White, Black, Hispanic, and “other”), employment status (employed, unemployed, student, and unable to work), education level (less than high school, high school graduate/General Educational Development exam completion, up to 3 years of college, and greater than 4 years of college), family yearly income (≤15,000, >15,000 to 25,000, >25,000 to 35,000, > 35,000-50,000, and > 50,000), marital status (married and unmarried), health insurance status (insured and uninsured), exercise reported in the past 30 days (yes, no), cigarette smoking status (current, former, never smoker), and history of ever being told to have asthma or chronic obstructive pulmonary disease.

### Statistical analysis

A descriptive analysis was conducted to assess overall sample characteristics and to check for missing data patterns. Subsequently, bivariate analyses were done to further assess for potential confounders in the sample. Lastly, unadjusted and adjusted logistic regression analysis were conducted to estimate crude and adjusted odds ratios and corresponding 95% confidence intervals to explore and control for potential confounding variables and to determine potential interactions (effect modifiers). STATA v15 software was used for all analyses.^11^ Further analyses restricted to participants who were never smokers and to those never reporting having asthma and COPD were performed.

This study does not require IRB clearance as it uses publicly available data from the CDC BRFSS.

## Results

Of the 22,844 participants who completed the BRFSS questionnaire in the selected states in 2017, 18,079 participants met our study inclusion criteria and were assessed. Table 1 displays the characteristics of participants according to e-cigarette use categories. Overall, never users of e-cigarettes were more frequently older, females, of minority race/ethnicity (African American and Hispanics), married, of higher education achievement (> 4 years of college) and higher income compared to all other e-cigarette users categories.

**Table 1:**
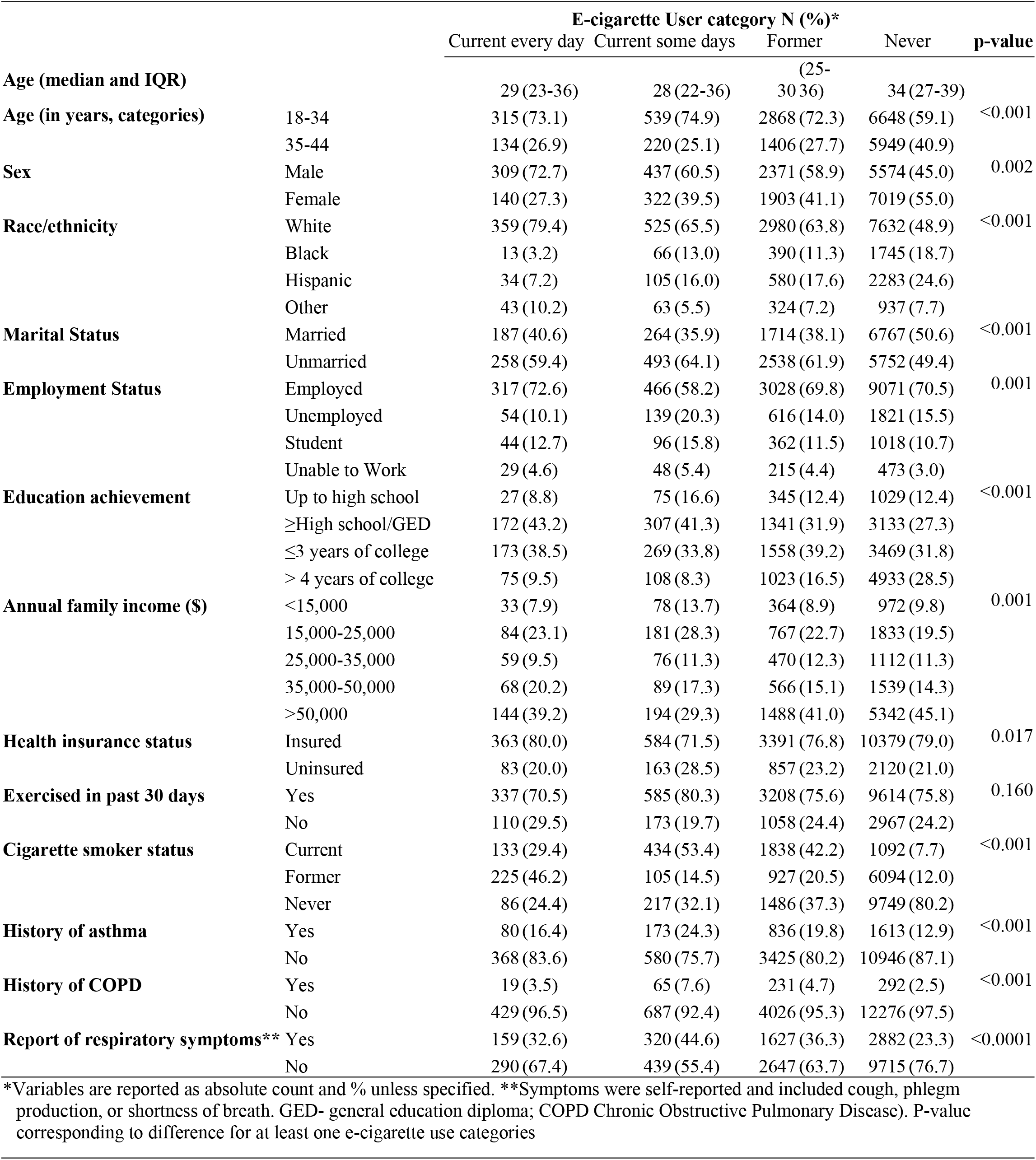
Selected characteristics of the sample of participants of the BRFSS according to e-cigarette use.

Never users also reported less frequently smoking of traditional (combustible) cigarettes. For most comparisons, differences were more striking between never users and everyday e-cigarette users. The frequency of asthma and COPD report were highest for those who e-cigarettes use frequency was reported as “some days”. The overall frequency of the respiratory symptoms assessed was 27.6%. This frequency varied according to e-cigarette use categorization: it was highest for those reporting as *someday current users* and lowest for *never users* of e-cigarettes (44.6% and 23.3%, respectively, p-value <0.0001 for differences in at least one category) (Table 1).

Table 2 displays the crude and adjusted odds ratios (ORs) for participant’s selected characteristics and reported respiratory symptoms. In the unadjusted analyses, compared to the never users, all e-cigarette users had significant increased odds of reporting respiratory symptoms. After controlling for confounding factors (participant’s demographic, socio-economic, behavioral -exercise and combustible cigarette use-asthma and COPD status), the associations were attenuated and significant only for those reporting some days of e-cigarette use and for former users compared to those who never used e-cigarettes (OR 1.49; 95% CI: 1.06-2.11and OR 1.29; 95% CI: 1.07-1.55, respectively). Other variables were also found independently associated with the occurrence of respiratory symptoms; Females, Blacks, those with lower education, being unable to work, family annual income lower than 50,000, being a current smoker, having no exercise reported in the past 30 days, and reporting asthma were associated with higher odds for respiratory symptoms in the adjusted analyses.

**Table 2.**
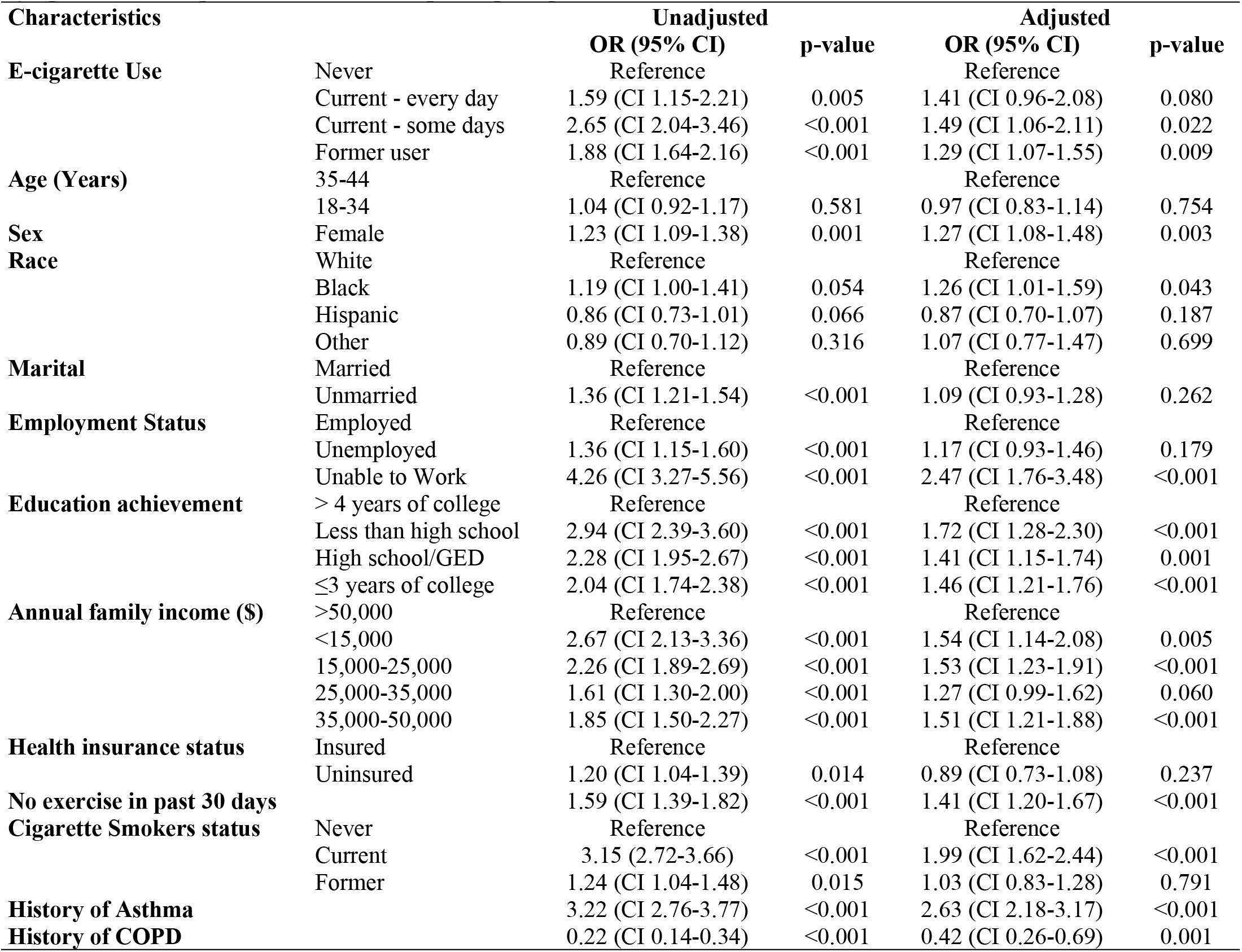
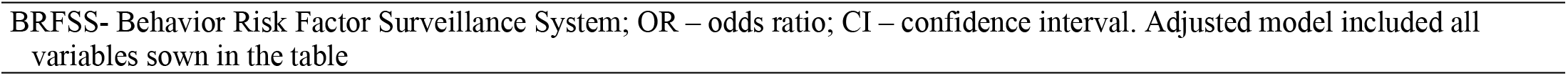
Associations between E-cigarette use, selected participant’s characteristics and occurrence of respiratory symptoms in a sample of adults in the US participating at the BRFSS 2017.

| Characteristics |  | Unadjusted |  | Adjusted |  |
| --- | --- | --- | --- | --- | --- |
|  |  | OR (95% CI) | p-value | OR (95% CI) | p-value |
| **E-cigarette Use** | Never | Reference |  | Reference |  |
|  | Current - every day | 1.59 (CI 1.15-2.21) | 0.005 | 1.41 (CI 0.96-2.08) | 0.080 |
|  | Current - some days | 2.65 (CI 2.04-3.46) | <0.001 | 1.49 (CI 1.06-2.11) | 0.022 |
|  | Former user | 1.88 (CI 1.64-2.16) | <0.001 | 1.29 (CI 1.07-1.55) | 0.009 |
| **Age (Years)** | 35-44 | Reference |  | Reference |  |
|  | 18-34 | 1.04 (CI 0.92-1.17) | 0.581 | 0.97 (CI 0.83-1.14) | 0.754 |
| **Sex** | Female | 1.23 (CI 1.09-1.38) | 0.001 | 1.27 (CI 1.08-1.48) | 0.003 |
| **Race** | White | Reference |  | Reference |  |
|  | Black | 1.19 (CI 1.00-1.41) | 0.054 | 1.26 (CI 1.01-1.59) | 0.043 |
|  | Hispanic | 0.86 (CI 0.73-1.01) | 0.066 | 0.87 (CI 0.70-1.07) | 0.187 |
|  | Other | 0.89 (CI 0.70-1.12) | 0.316 | 1.07 (CI 0.77-1.47) | 0.699 |
| **Marital** | Married | Reference |  | Reference |  |
|  | Unmarried | 1.36 (CI 1.21-1.54) | <0.001 | 1.09 (CI 0.93-1.28) | 0.262 |
| **Employment Status** | Employed | Reference |  | Reference |  |
|  | Unemployed | 1.36 (CI 1.15-1.60) | <0.001 | 1.17 (CI 0.93-1.46) | 0.179 |
|  | Unable to Work | 4.26 (CI 3.27-5.56) | <0.001 | 2.47 (CI 1.76-3.48) | <0.001 |
| **Education achievement** | > 4 years of college | Reference |  | Reference |  |
|  | Less than high school | 2.94 (CI 2.39-3.60) | <0.001 | 1.72 (CI 1.28-2.30) | <0.001 |
|  | High school/GED | 2.28 (CI 1.95-2.67) | <0.001 | 1.41 (CI 1.15-1.74) | 0.001 |
|  | ≤3 years of college | 2.04 (CI 1.74-2.38) | <0.001 | 1.46 (CI 1.21-1.76) | <0.001 |
| **Annual family income (\$)** | >50,000 | Reference | | Reference | |
|  | <15,000 | 2.67 (CI 2.13-3.36) | <0.001 | 1.54 (CI 1.14-2.08) | 0.005 |
|  | 15,000-25,000 | 2.26 (CI 1.89-2.69) | <0.001 | 1.53 (CI 1.23-1.91) | <0.001 |
|  | 25,000-35,000 | 1.61 (CI 1.30-2.00) | <0.001 | 1.27 (CI 0.99-1.62) | 0.060 |
|  | 35,000-50,000 | 1.85 (CI 1.50-2.27) | <0.001 | 1.51 (CI 1.21-1.88) | <0.001 |
| **Health insurance status** | Insured | Reference |  | Reference |  |
|  | Uninsured | 1.20 (CI 1.04-1.39) | 0.014 | 0.89 (CI 0.73-1.08) | 0.237 |
| **No exercise in past 30 days** |  | 1.59 (CI 1.39-1.82) | <0.001 | 1.41 (CI 1.20-1.67) | <0.001 |
| **Cigarette Smokers status** | Never | Reference |  | Reference |  |
|  | Current | 3.15 (2.72-3.66) | <0.001 | 1.99 (CI 1.62-2.44) | <0.001 |
|  | Former | 1.24 (CI 1.04-1.48) | 0.015 | 1.03 (CI 0.83-1.28) | 0.791 |
| **History of Asthma** |  | 3.22 (CI 2.76-3.77) | <0.001 | 2.63 (CI 2.18-3.17) | <0.001 |
| **History of COPD** |  | 0.22 (CI 0.14-0.34) | <0.001 | 0.42 (CI 0.26-0.69) | 0.001 |

To study potential interactions, further analyses were conducted assessing the association between e-cigarette use, and respiratory symptoms occurrence in participants who were never smokers. The absolute number of participants who never smoked combustible cigarettes and were 86, 217, 1486, and 9749 for current every day, current some days, former, and never e-cigarette users, respectively. In this analysis, the only significant increased odds for respiratory symptoms were found for former e-cigarette users compared to never users (supplemental table 1). Lastly, exclusion of participants with prior history of asthma and COPD did not affect the estimates of the associations reported for e-cigarette and respiratory symptoms compared to the analyses where those conditions were adjusted for (supplemental table 2).

## Discussion

Our data revealed that 2.5% and 4.2% of the sample subjects were *every day* or *some days* e-cigarette users, respectively. Overall, respiratory symptoms (cough, phlegm production, or shortness of breath) were reported in approximately 28% of the sample, a much higher frequency than those symptoms previously reported in another research. (9-11) Compared to never users, e-cigarette use was associated with up to 50% increased odds of respiratory symptoms; the highest odds were found for those using some day, followed by former users of e-cigarette.

While previous research demonstrated that even short-term exposure to e-cigarettes and their toxic compounds produced cough and phlegm in healthy individuals and individuals with a history of asthma,^7^ most of the previous cross-sectional and longitudinal studies in US adults that were published, assessed the detrimental effects of e-cigarettes focusing on outcomes such as asthma and COPD occurrence (12,13,15,17). Our results for someday e-cigarette users suggesting a 49% increase in odds for the outcome, a composite of cough, phlegm production, or shortness of breath, were consistent in magnitude with the previous studies, despite assessing associations independently of COPD, asthma status, and demographic, socio-economic, and clinical characteristics. For instance, in the meta-analyses reported in the integrative review published in 2021 by Wills et al, the pooled odds of asthma and COPD were 39% (OR=1.39, 95% CI 1.28-1.51) and 49% (OR1.49, 95% CI=1.36-1.65) higher, respectively, for e-cigarette users compared to never users (12). A less marked increase in the odds of cough or phlegm has also been reported in Chinese adolescents in 2012-2013, with e-cigarette users having 1.28 times higher odds to develop symptoms compared to non-users (regardless of other combustible smoking habits). (11) Our results suggest that increases symptoms occur, and are independent of asthma and COPD diagnosis. We also attempted to further assess for potential gradient in the risk according to the frequency of e-cigarette use and our findings indicate that - while for current everyday users the magnitude of the association seemed like the one found for some day users - results were no longer significant after adjustments. It is possible that the smaller number of participants (about 40% less than some days users) might have contributed to the lack of statistical significance. Prospective studies including larger number of e-cigarette users are needed to better understand causality better, and to define whether a dose-response gradient exists for the risk of e-cigarettes use and respiratory symptoms.

E-cigarettes are designed to deliver nicotine throughout the body, without the combustion of tobacco. Through inhalation of water vapor to the lungs, numerous toxic compounds like those found within conventional cigarettes are released within the body. Examples of these compounds are aldehydes (formaldehyde and acrolein) and e-liquids (propylene glycol and glycerol). (7,8,18) Overtime, these chemicals cause an elevated mucin concentration and result in failed mucus transport, hallmarks seen in patients of COPD (Chronic Obstructive Pulmonary Disease). The chemicals within e-cigarettes also stimulate neutrophils, resulting in the release of myeloperoxidase (MPO), neutrophil elastase (NE), and other granular proteins. The amount of proteins released in e-cigarettes is double to what was described for conventional cigarettes, and it causes massive structural damage to airways. (19) These changes could explain potential mechanism that contribute to the common symptoms of chronic cough, phlegm, and shortness of breath observed in e-cigarette users.

This study large sample size and the use of systematically collected and comprehensive information from participants from multiple states contribute to increase study’s generalizability when representing the adult population in the US. There are some limitations inherent to the cross-sectional and self-reported nature of the survey, and they should be discussed. First, we lack detailed information on the timing when e-cigarette and the symptoms occurred. The BRFSS survey inquired about cough and phlegm occurring in the three months preceding the survey, while e-cigarettes use status referred to the time of the survey. Therefore, the extent of which potential reverse causality accounts for the association described is unknown, and prospective studies are warranted. Furthermore, given the responses were based on self-report, misclassifications of both exposure to e-cigarettes and occurrence of symptoms are possible. While we do not have evidence to help us to estimate the directionality of the bias, we believe that both the frequency of symptoms and of e-cigarettes would be underreported, thus, most likely biasing the association towards the null hypothesis.

In conclusion, our results also suggest that e-cigarettes might increase the risk of detrimental respiratory symptoms, but the dose-effect relationship is yet to be confirmed with more accurate assessment of e-cigarette exposure levels. Therefore, our study adds to the growing body of literature suggesting that e-cigarettes are not safe alternatives for smoking cessation interventions.

## Data Availability

Data for the study is available publicly at the CDC BRFSS site:https://www.cdc.gov/brfss/annual_data/annual_data.htm

## Acknowledgments

We thank the Behavior Risk Factor Surveillance System (BRFSS) working group and the survey participants for their contribution.

## Notes

### Competing Interest Statement

The authors have declared no competing interest.

### Funding Statement

This study is an independent study and has not received any sponsorship, is not part of a grant, or has received any funding.

### Author Declarations

Non human subjects research (publicly available anonymous data)

